# Beyond Education and Contraceptive Use: Do Caste, Ethnicity, and Religion Influence Fertility Behavior in Nepal? Evidence from Six Rounds of DHS

**DOI:** 10.1101/2025.09.03.25335049

**Authors:** Dhurba Ghimire, KC Samir

## Abstract

**Background:** Nepal has experienced a significant decline in fertility rates. However, notable variations persist, necessitating further analysis. This study systematically examines the determinants of fertility heterogeneity in Nepal, focusing on caste, ethnicity, religion, and related factors.

**Methods:** The study analysed data from six rounds of the Nepal Demographic and Health Survey (1996–2022) to examine married women aged 15-49. A Poisson regression model was used to assess the relationship between various determinants and the Number of Children Ever Born (CEB).

**Results:** Significant associations (p < 0.001) with CEB were identified. Women marrying at age ≥20 experienced a 25% reduction in expected CEB compared to those marrying at ≤19. Those with a met need for contraception had a 9.2% lower CEB, while urban women exhibited an 11.1% reduction compared to rural women. Muslim women had an 11.1% higher CEB, whereas Dalit and Janajati women showed lower CEB compared to Arya women. Additionally, women from poorer households had a 9.2% lower CEB, and educational attainment significantly influenced CEB, with post-secondary education linked to a 48.5% reduction in CEB.

**Conclusion:** The study reveals some level of heterogeneity in fertility rates across caste, ethnicity, and religion. It emphasises the importance of female education, family planning, and women’s empowerment in reducing fertility rates and suggests that targeted government investments can address these disparities.

## Background

The principal components of a population change in any country are fertility, mortality, and migration. Among these, fertility is the most significant factor influencing population growth, structure, and composition (1). Historically, reproduction increased when resources were abundant for all kinds of species, including plants, animals, and humans, before the Industrial Revolution (2). During that era, households with higher incomes had more surviving children than those with lower incomes(3). However, economic growth led to a demographic transition from high fertility to low fertility (4). Today, both within and across countries, wealthier and more educated households tend to have fewer children than poorer and less educated ones (5,6). This paper focuses on fertility and its heterogeneity, defined as multiple trajectories within a cohort of women that are significantly different from one another(7).

Research on fertility decline has primarily focused on family planning interventions and socioeconomic development in developing countries(8–11). Many Low- and Middle-Income Countries (LMICs) in South Asia and Sub-Saharan Africa have experienced rapid declines in fertility (12–14). Nepal, being an LMIC (15), succeeded in lowering its total fertility rate to replacement level over the last few decades, from 6.3 in 1981 (16) to 2.1 in 2022 (17).

Previous studies on proximate determinants of fertility in Nepal have used aggregate modelling techniques, focusing on four major factors: (a) proportion of women married or in sexual union or total marital fertility, (b) contraceptive use and estimated effectiveness, (c) duration of postpartum infecundability, and (d) induced abortion (18–21) . Other contextual factors that affect fertility include women’s education and employment, women’s empowerment, urbanisation, societal prosperity, desired family size, and access to reproductive health services (20). These individual and contextual factors are interrelated and reinforce each other (22–24).

Educational attainment of women is negatively correlated with fertility (25–27). Higher education among reproductive women is associated with increased contraceptive use, better public health care utilisation, and a better understanding of child-rearing costs, leading to lower fertility (28,29). Education also enhances human capital, providing women with knowledge, skills, and opportunities that often conflict with repeated childbearing (30,31). Additionally, formal education provides women with gainful employment, increasing labour force participation and limiting childbearing (32,33).

Education also raises the mean age at marriage, enhances women’s decision-making roles, and promotes smaller family sizes (34–41) It improves knowledge of family planning and increases the opportunity costs of childbirth (42). Family planning programmes have effectively increased access to contraception and promoted smaller families, or often do both. These studies are mainly in Asian countries with rapid fertility declines, such as Bangladesh, Indonesia, Iran, and Nepal (43–47).

While the literature highlights the impact of education on fertility, the interplay of education with various covariates remains crucial. This study examines the influence of education on fertility and its connection with factors such as age at first marriage, place of residence, caste/ethnicity dynamics, unmet need for contraception, and women’s empowerment. Fertility levels in Nepal vary beyond the urban-rural context and encompass variations in educational attainment.

Caste and ethnicity significantly shape Nepal’s social structure. The caste system comprises four major groups: Brahmin, Kshetri, Vaishya and Shudra. Additionally, ethnic diversity in Nepal includes groups such as Sherpa, Tamang, Magar, Gurung, Rai, Limbu, and Tharu. These groups contribute significantly to Nepal’s cultural richness, with historical, social, and economic factors all influencing its social dynamics. In recent decades, caste/ethnicity have been focal points in research, serving as lenses to examine disparities in socioeconomic development, particularly in areas such as poverty, education, and health (48–52).

Studies show persistent caste and ethnic differentials in fertility. For example, The NDHS 2022 reports the highest TFR among Muslim groups (3.3 children per woman), followed by Madheshi and Dalit communities (2.4 each), and the lowest among Janajati (1.8). The major cause of these differentials is rooted in historical, social, and cultural discrimination, which impacts health outcomes (48,53,54).

There is limited evidence on fertility heterogeneity among women in terms of caste/ethnicity, education, and associated covariates. This study examines Nepal’s heterogeneous fertility structure by analysing women’s fertility trajectories across the six rounds of the NDHS (1996-2022). It aims to determine the magnitude and determinants of fertility heterogeneity among Nepali women, with an emphasis on fertility differentials by caste and ethnicity, as well as other associated covariates. Additionally, this paper seeks to guide demographers and planners in projecting future populations, as fertility is a vital component of population change.

### Conceptual Framework

The conceptual framework for determining the determinants of fertility heterogeneity in Nepal is based on an extensive literature review, aligning with the study’s objectives. Several studies have shown that women’s fertility behaviours are influenced by several factors (18,25,31,39,55–60). These factors include the age of women, recognising the impact of maternal age on fertility outcome; age at first marriage, acknowledging the significance of the timing of marriage concerning fertility; marital status, considering the influence of marital status on reproductive decisions; caste/ethnicity, addressing the sociocultural dimensions tied to fertility variations among different ethnic groups; place of residence, exploring distinct fertility patterns between urban and rural settings; household wealth, understanding the economic factors affecting fertility; educational attainment, emphasising the pivotal role of education in shaping social norms and women’s empowerment; and contraceptive use, analysing the impact of family planning methods on fertility outcomes.

In this study, education is highlighted as a catalyst for changing social norms and empowering women. The conceptual framework visualises the intricate interplay of these variables, illustrating their collective influence on fertility outcomes, as presented in **Fig 1**.

**Fig 1.**
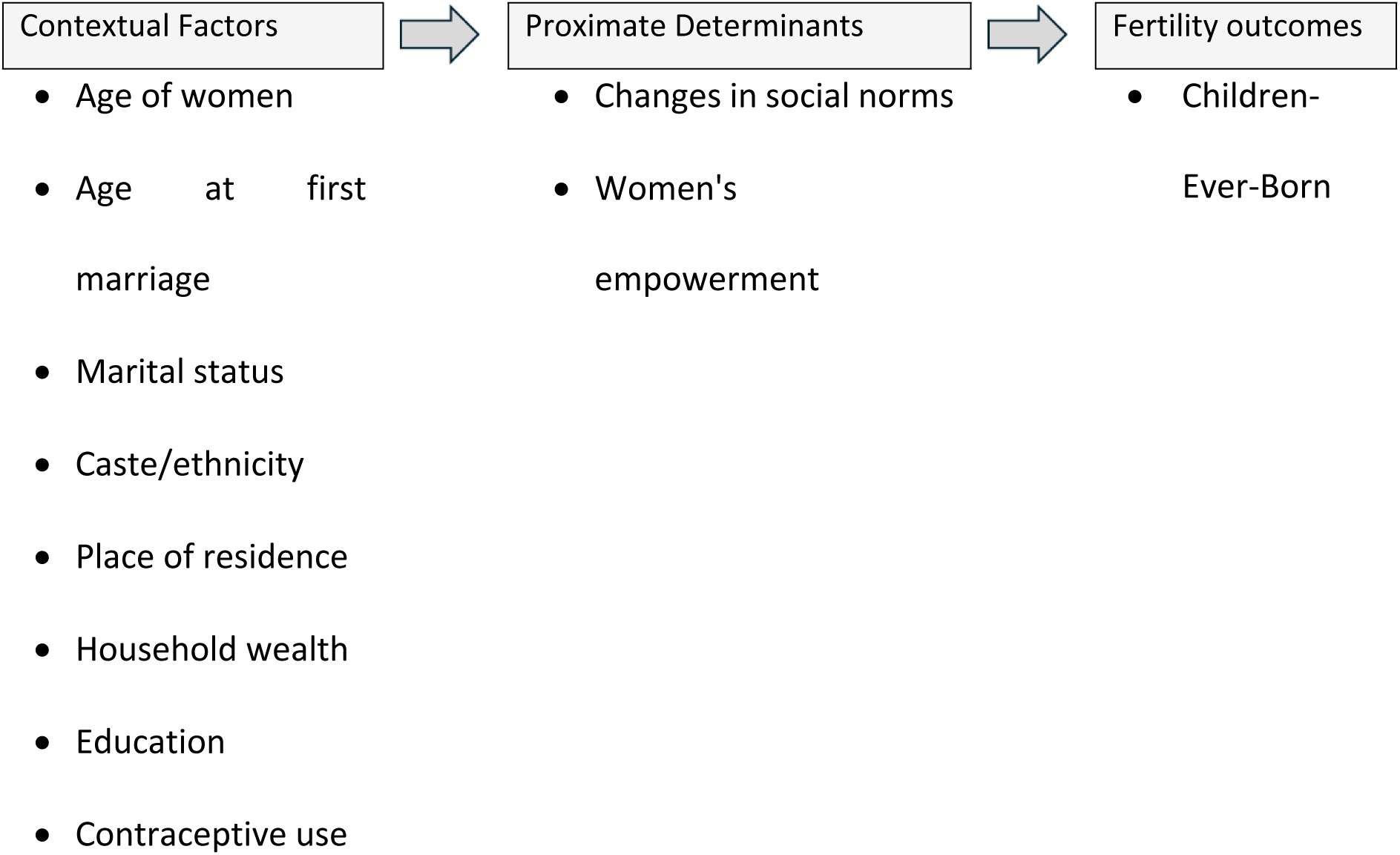
Conceptual Framework of Heterogeneity of Fertility

## Methods and Materials

### Study Design, Population and Period

The study utilised six rounds of the Nepal Demographic Health Survey (NDHS) from 1996, 2001, 2006, 2011, 2016, and 2022. The NDHS is a nationally representative household survey conducted under the authority of the Ministry of Health and Population, Government of Nepal, and was implemented by the NGO New Era in Kathmandu, Nepal. The datasets were retrieved from the Demographic Health Survey (DHS) program’s official website (http://dhsprogram.com). DHS datasets are collected using a stratified, multi-stage cluster random sampling design. It is a part of the worldwide DHS programme, which collects data on fertility, family planning, and maternal and child health.

Details on the sample design and survey methodology can be found in the respective NDHS reports. Individual-level datasets are available from the DHS data repository and can be accessed upon request (61). The target population for this study was currently married women of reproductive age at the time of the survey. This generated a sub-sample of 55,168 currently married women from a pooled dataset of 57,590 ever-married women of reproductive age.

Table 1 provides the sample sizes for the different rounds of the NDHS implemented in Nepal.

**Table 1.**
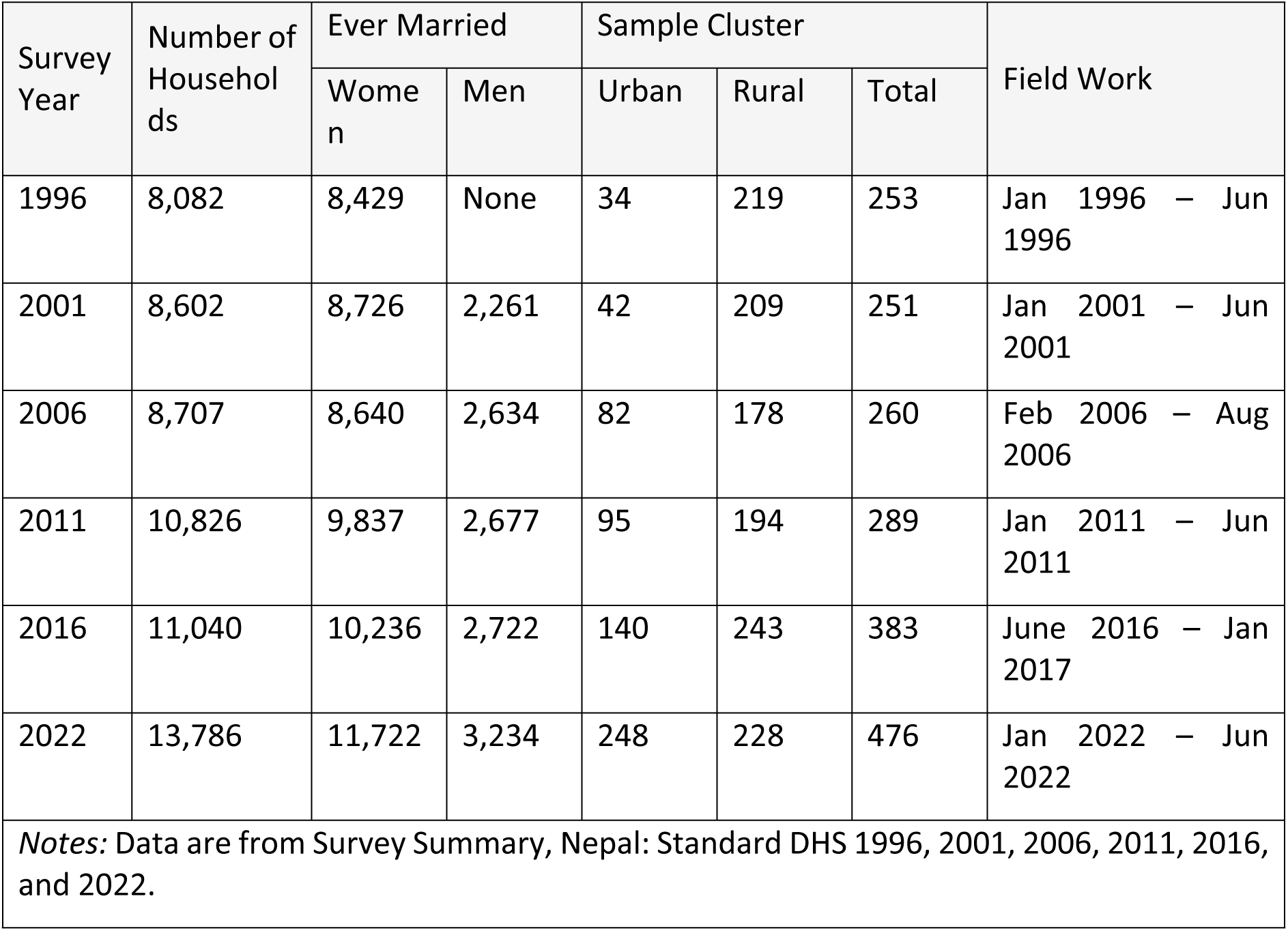
Sample coverage of the Nepal Demographic Health Survey (NDHS) in 1996-2022.

| Survey Year | Number of Households | Ever Married |  | Sample Cluster |  |  | Field Work |
| --- | --- | --- | --- | --- | --- | --- | --- |
|  |  | Women | Men | Urban | Rural | Total |  |
| 1996 | 8,082 | 8,429 | None | 34 | 219 | 253 | Jan 1996 – Jun 1996 |
| 2001 | 8,602 | 8,726 | 2,261 | 42 | 209 | 251 | Jan 2001 – Jun 2001 |
| 2006 | 8,707 | 8,640 | 2,634 | 82 | 178 | 260 | Feb 2006 – Aug 2006 |
| 2011 | 10,826 | 9,837 | 2,677 | 95 | 194 | 289 | Jan 2011 – Jun 2011 |
| 2016 | 11,040 | 10,236 | 2,722 | 140 | 243 | 383 | June 2016 – Jan 2017 |
| 2022 | 13,786 | 11,722 | 3,234 | 248 | 228 | 476 | Jan 2022 – Jun 2022 |
| *Notes:* Data are from Survey Summary, Nepal: Standard DHS 1996, 2001, 2006, 2011, 2016, and 2022. |  |  |  |  |  |  |  |

### Study Variables

#### Outcome Variable

The outcome variable, Children Ever Born (CEB), refers to the number of children born alive to a woman during her lifetime and serves as a measure of lifetime fertility (62). The mean number of CEB to women represents the childbearing experience of a real-age cohort and reflects both current and past fertility behaviour. CEB enables the generalisation of data and understanding of fertility, providing a basis for further analysis (63). The CEB for all women, irrespective of age group (lifetime fertility), and for women aged 40-49 (completed fertility) were analysed separately as a measure of fertility.

#### Predictor Variables

The demographic, socioeconomic and sociocultural factors considered as predictor variables in this study include (a) age of respondents, (b) age at first marriage, (c) place of residence, (d) caste/ethnicity, (e) educational attainment, (f) household wealth status, (g) unmet need of family planning and (h) women’s empowerment. These predictor variables were selected for inclusion in the analysis based on a comprehensive literature review on fertility (64–66) and their hypothesised association with CEB and significance in previous studies (20,25,55,57,67). The variables have been categorised conventionally, as used in other studies, and based on their frequency distribution. To make the analysis and interpretation more meaningful and comparable, some variables were regrouped from their original categories in the dataset.

The following paragraph summarises the categorisation of the study variables.

- Age at first marriage: Categorised into adolescents (≤ 19 years) and non-adolescents (≥ 20 Years).
- Place of residence: Urbanisation comparisons over time in Nepal are subject to limitations due to political decisions that lack transparent and standardised criteria for declaring urban areas. The Constitution of Nepal (2015) defines urban areas as metropolitan, sub-metropolitan, municipality and rural municipalities. NDHS surveys follow the Government of Nepal’s definitions. However, results should be interpreted cautiously due to inconsistencies and a lack of standardisation of the specific thresholds and factors considered over time.
- Caste/ethnicity: The term “caste” refers to hierarchical groups within Hindu religious values of purity and impurity (68), whereas “ethnicity” encompasses cultural attributes such as a collective name, shared history, and association with a specific territory (69). Nepal’s Constitution (2015) categorises caste/ethnic groups into Khas/Arya, Janajati, Madheshi, Dalit and Muslim. The Khas/Arya, referred to as Arya, comprises castes such as Kshetri, Brahmin, Thakuri, and Sanyasi (70). The Janajati group includes 60 ethnicities (71). Similarly, the Madheshi community comprises 150 castes and ethnicities (72), and Dalits include 26 castes (73).
- Educational attainment: Nepal’s education system comprises preschool, basic (grades 1-8), secondary (grades 9-12), and tertiary (bachelor’s, master’s, and PhD) levels (74). According to the International Standard Classification of Education, 2011 (75), educational attainment ranges from level 0 (early childhood education) to level 8 (doctoral or equivalent). To maintain consistency across NDHS rounds, educational attainment has been classified into six groups equivalent to ISCED levels: no education, incomplete primary (level 0), primary (level 1), lower secondary (level 2), upper secondary (level 3), and post-secondary (level 4-8).
- Household wealth status: Categorised into five groups: poorest, poorer, middle, richer, and richest.
- Unmet needs for family planning: Categorised into met need and unmet need.
- Women’s empowerment: This variable is derived from decision-making in four areas: healthcare, spending one’s earnings, visits to family or relatives, and large household purchases. It is coded as follows: 0 (no decision-making), 1 (decision-making in at least one area), and 2 (decision-making in all four areas).

### Data Management and Method of Statistical Analysis

To examine the factors influencing fertility, three statistical approaches were used. First, descriptive univariate analysis was used to inspect the frequency distribution of the study variables. Second, bivariate analysis examined the relationships between predictor variables and the CEB. Since CEB is continuous, the relationship between the mean number of CEB and the predictor variables was analysed using one-way ANOVA and an F-test to determine fertility differentials.

Lastly, the effect of the main explanatory variables and covariates on the outcome variable was analysed using two Poisson regression models, including all control variables. Model 1 measured the net effect of each variable on lifetime fertility among women aged 15-49. Model 2 measured the net effect on completed fertility among women aged 40-49.

#### Analysis of Variance (ANOVA)

In performing the ANOVA, the study considered the assumptions of independence of observations, normal distribution, and homogeneity of variances (76,77). The assumption of independent observation was met, as DHS surveys ensured that each woman was interviewed only once. The assumption of normal distribution was not met, as the Kolmogorov-Smirnov normality test on the outcome variable showed a p-value significantly less than 0.5, thus suggesting a significant deviation from normality. However, the large sample size was sufficient for the use of ANOVA, even though the normal distribution was lacking (78).

The test for homogeneity of variances was performed using robust methods (79,80), which assesses the null hypothesis that variances across groups are equal. This assumption, crucial for t-tests and F-tests (ANOVA) (81), was not met according to Levene’s test. Alternative F statistics (Welch’s or Brown-Forsythe) were used to determine statistical significance (82). Subsequently, a one-way ANOVA was used to analyse the association between the mean number of CEB and selected predictor variables.

Post-hoc tests, including Temhane, Dunnett T3 and Games-Howel (77), were conducted due to failure to meet the equal variance assumption. These tests, which compare group means after ANOVA produced almost identical results. Given ANOVA’s limitation to one independent variable, a multivariable Poisson regression model was used to estimate the net effect of women’s autonomy on fertility, controlling for other covariates.

#### Poisson Regression Model

Poisson’s distribution – a limiting form of binomial distribution, is suitable for discrete/count outcome variables (83–86). Thus, the Poisson regression model is appropriate for the count outcome variable, CEB. The Poisson regression model is expressed as (86),

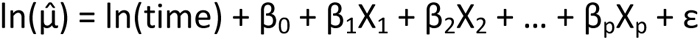

where 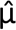 is the predicted count of the outcome variable given the predictors X_1_, X_2_, …, X_p_. Where ln refers to the natural logarithm, β_0_ is the intercept, β_1_ is the regression coefficient for the first predictor X_1_, and ln(time) represents an offset variable. The Poisson error structure (ε) resolves issues with applying Ordinary Least Squares (OLS) regression to count outcomes, such as non-constant variance and the non-normal conditional distribution of errors. Coefficients were exponentiated to yield the Incident Rate Ratio (IRR) for easier interpretation of the results. The IRR explains how a change in X affects the rate of the outcome variable (85). Thus, the results are presented and interpreted as IRR with a 95% Wald confidence interval (WCI) (62).

The assumption of equal variance in the Poisson regression model was tested using Akaike’s Information Criterion (AIC) and Bayesian Information Criterion (BIC). If this assumption is violated, the negative binomial model is recommended (87–89). This study found under-dispersion (the conditional variance is less than the predicted mean), but over-dispersion (the conditional variance is greater than the predicted mean) is more concerning (90). A Negative Binomial model was also applied to correct under-dispersion. The best fit model, determined by lower AIC and BIC (89), was the Poisson regression model, which produced lower indices.

The Poisson regression model is applicable when counts are made within a fixed time interval. Where counts vary across cases (e.g., age of the mother), an *exposure variable* – a covariate with a fixed coefficient of 1 - adjusts for differences in the observation period, converting raw counts to rates (83). This study used the *current age of women as the exposure variable*, incorporated as a natural log (91).

The DHS sample, including the NDHS, is weighted and employs a complex sample design involving stratification and clustering. Weighting variables were used in complex sample analysis procedures for descriptive analysis. There is debate about using weights in multivariate models, but the consensus is to use them for descriptive statistics (92–95). This study applied complex sample analysis for both descriptive and inferential statistics.

Analysis was performed separately for all women and women aged 40-49 in both bivariate and multivariate analyses. A correlation matrix was used to assess the relationship between the outcome variable and predictor variables. The results demonstrated that there was no multicollinearity (*r > 0.6*) between predictor and outcome variables. All the variables from the bivariate analysis were included in the multivariate Poisson regression model. The regression model was performed using Stata software version 18. The svyset command in Stata was used to estimate the correct standard errors, adjusted for stratification, clustering, and weights.

## Results and Discussions

### Characteristics of Respondents

The total weighted sample of 55,243 currently married women of reproductive age (15-49 years) was used for this study, drawn from the individual and pooled NDHS datasets. Table 2 presents the mean and variance of children ever born across six survey rounds. Over the study period, the mean number of children decreased from 3.41 in 1996 to 2.81 in 2022. For women aged 40-49, the rate declined from 5.93 in 1996 to 3.31 in 2022. The observed variance for each year and pooled estimates reveal that the variance, which should not exceed twice the mean value, does not deviate significantly from the mean, thus supporting the assumption of the Poisson distribution.

**Table 2.** Mean and variance of children ever born, NDHS (1996–2022)

| Age Group | 1996 |  | 2001 |  | 2006 |  | 2011 |  | 2016 |  | 2022 |  | Pooled |  |
| --- | --- | --- | --- | --- | --- | --- | --- | --- | --- | --- | --- | --- | --- | --- |
|  | Mean | Var | Mean | Var | Mean | Var | Mean | Var | Mean | Var | Mean | Var | Mean | Var |
| 15–49 Years | 3.41 | 6.24 | 3.29 | 5.63 | 3.04 | 4.72 | 2.68 | 3.63 | 2.48 | 3.06 | 2.25 | 2.22 | 2.81 | 4.27 |
| 40–49 Years | 5.93 | 6.60 | 5.60 | 6.08 | 5.06 | 5.03 | 4.36 | 4.37 | 3.83 | 3.54 | 3.31 | 2.63 | 4.50 | 5.32 |
| Notes: N = Weighted N. Data are from the author's computation from the pooled six-round NDHS dataset (1996 – 2022) |  |  |  |  |  |  |  |  |  |  |  |  |  |  |

Table 3 provides the background characteristics of the respondents. More than three-fourths of the respondents were married by the time they were 19 years old or younger throughout the study period (1996–2022). The majority (> 85%) lived in rural areas from 1996 to 2011, but this dropped to 38.9% in the 2016 survey and 32.4% in 2022. This change was due to the Government of Nepal’s official declaration of new urban areas and the expansion of existing urban areas.

Regarding caste and ethnicity, Janajati remained the largest group, comprising over one-third of the respondents. Arya was the second-largest group, accounting for around 28%, while the Dalit and Madheshi communities each comprised approximately 15%, and Muslims made up around 5%. Educational attainment improved significantly, with the share of respondents with no formal education decreasing from 81% in the 1996 survey to 32% in the 2022 survey.

Table 3 also reveals that 21.3% of the respondents were in the poorest wealth quintile, while 19.2%, 19.4%, 19.9% and 20.3% were in the poorer, middle, richer and richest wealth quintiles, respectively. More than two-thirds of respondents had their family planning needs met throughout the study period, while the share of unmet needs decreased from 31.4% in 1996 to 25.6% in 2022. Regarding women’s empowerment, over two-thirds (64.4%) of respondents lacked decision-making capacity, followed by 32.5% with partial decision-making capacity and 3.1% with full decision-making capacity throughout the study period.

**Table 3.**
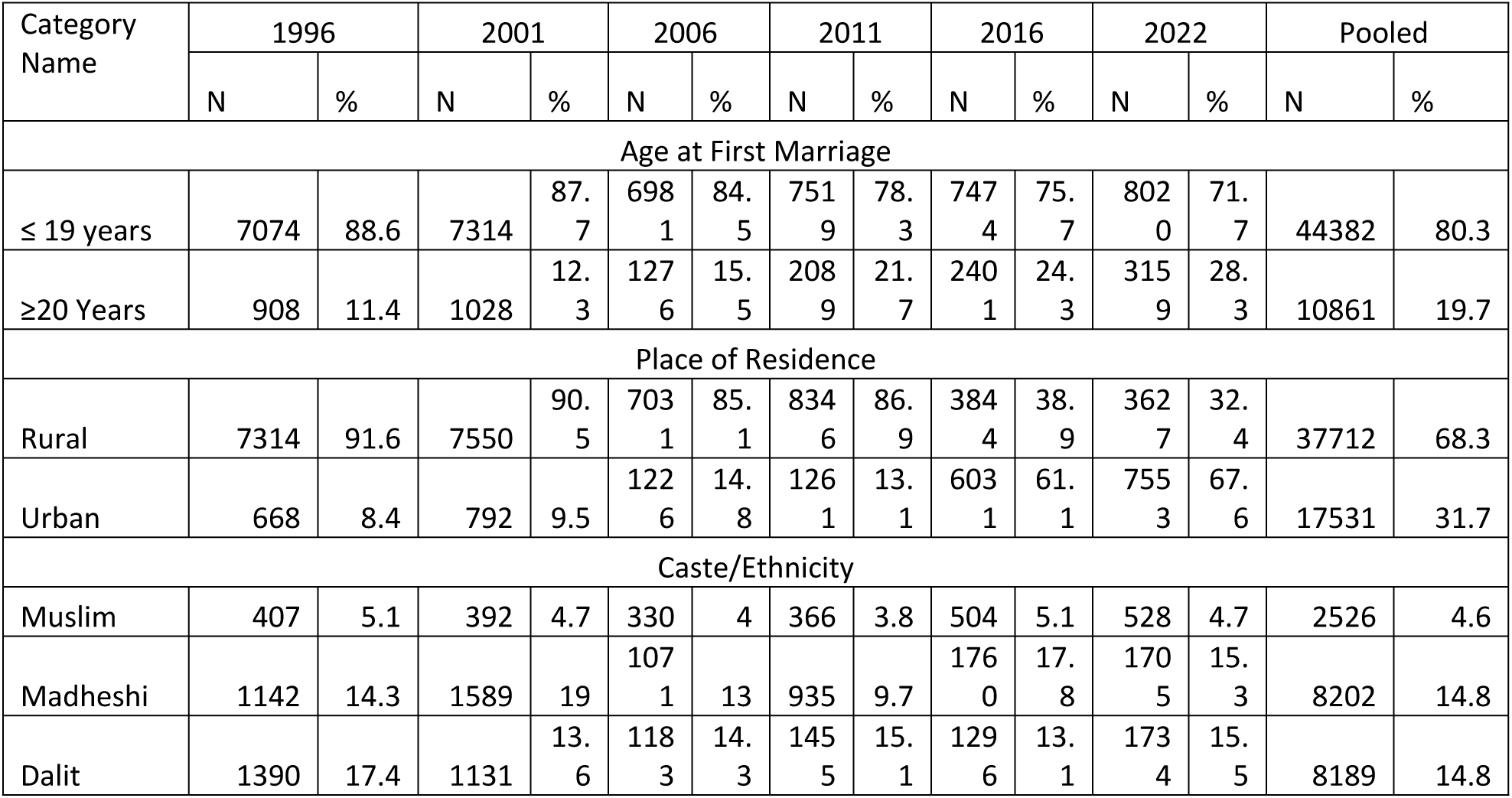
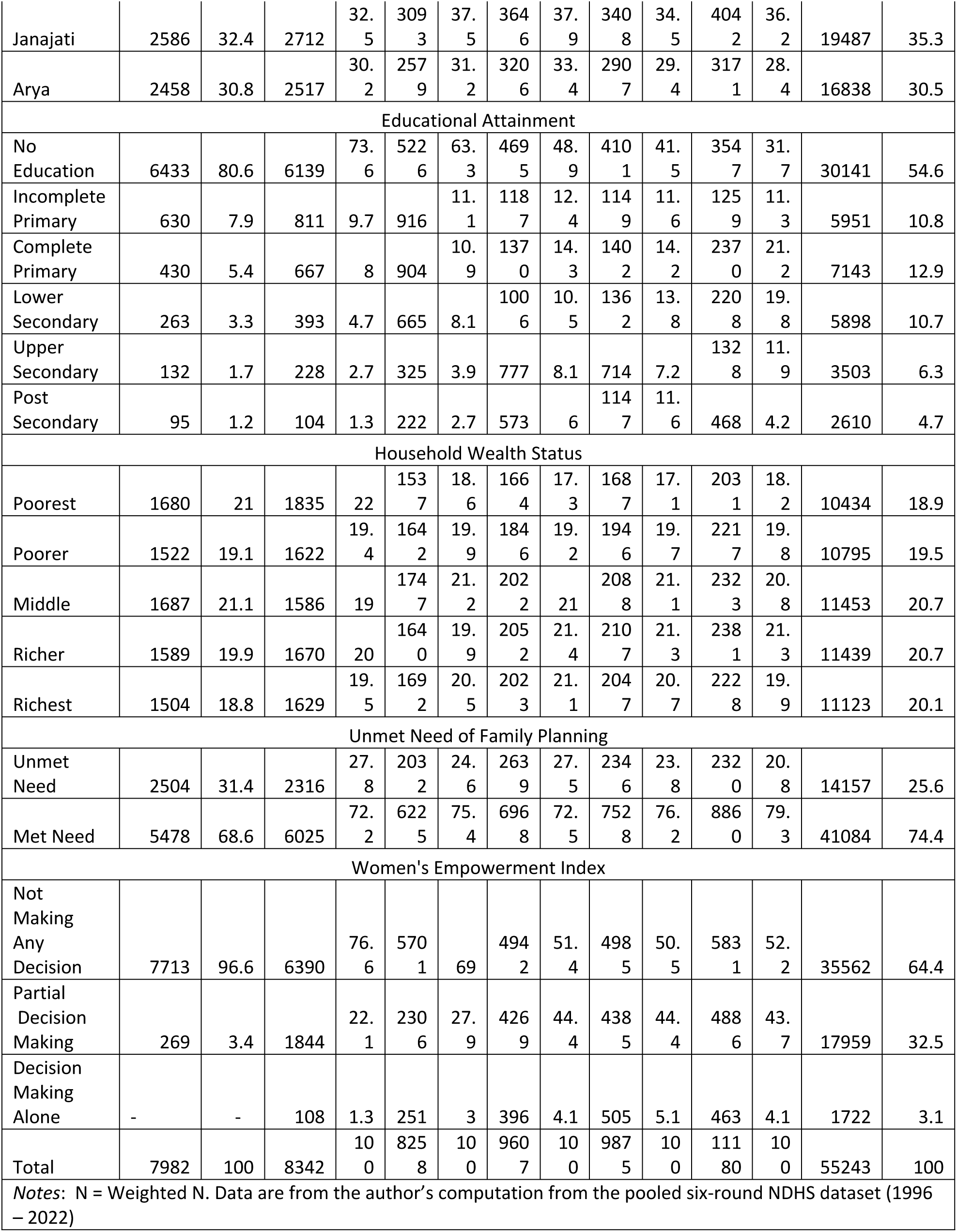
Distribution of currently married women of reproductive age (15-49 years) by selected characteristics at the time of the survey, NDHS (1996–2022).

### Bivariate Analysis

The association between CEB and each predictor variable for two categories – women of reproductive age (15-49 years) and those in the completed fertility age group (40–49 years) – was analysed using one-way ANOVA. The results are summarised in Table 4. The statistical analysis revealed significant associations between the number of CEB and the following variables: age at first marriage, usual place of residence, caste/ethnicity, educational attainment, household wealth status, unmet need of family planning, and women’s empowerment index. Consequently, all significant control variables were included for multivariable Poisson regression analysis.

**Table 4.** Bivariate Analysis (Based on ANOVA) of Individual Variables with Total Number of Children Ever Born (CEB) with Currently Married Women of Reproductive Age, NDHS (1996–2022)

| Categories | All Women (15-49 years) |  |  |  | Women Aged 40 - 49 |  |  |  |
| --- | --- | --- | --- | --- | --- | --- | --- | --- |
|  | Mean CEB | SD | 95% CI | F-Score | Mean CEB | SD | 95% CI | F-Score |
| Age at First Marriage |  |  |  |  |  |  |  |  |
| ≤ 19 years | 3.02 | 2.1 | 2.98 - 3.05 | 2327.8*** | 4.79 | 2.3 | 4.71 - 4.88 | 818.2*** |
| ≥ 20 years | 1.97 | 1.6 | 1.92 - 2.01 |  | 3.33 | 1.9 | 3.23 - 3.44 |  |
| Place of Residence |  |  |  |  |  |  |  |  |
| Rural | 3.05 | 3.1 | 3.01 - 3.09 | 1662.0*** | 5.05 | 2.4 | 4.96 - 5.14 | 1475.8*** |
| Urban | 2.29 | 2.3 | 2.24 - 2.34 |  | 3.43 | 1.8 | 3.33 - 3.54 |  |
| Caste/Ethnicity |  |  |  |  |  |  |  |  |
| Muslim | 3.45 | 2.5 | 3.29 - 3.6 | 119.2*** | 6.2 | 2.6 | 5.86 - 6.55 | 115.6*** |
| Madheshi | 2.97 | 2 | 2.91 - 3.03 |  | 4.62 | 2 | 4.48 - 4.76 |  |
| Dalit | 2.99 | 2.2 | 2.92 - 3.06 |  | 5.06 | 2.3 | 4.9 - 5.23 |  |
| Janajati | 2.68 | 2.1 | 2.62 - 2.75 |  | 4.34 | 2.4 | 4.21 - 4.48 |  |
| Arya | 2.69 | 2 | 2.65 - 2.74 |  | 4.2 | 2.2 | 4.09 - 4.32 |  |
| Educational Attainment |  |  |  |  |  |  |  |  |
| No Education | 3.62 | 2.2 | 3.58 - 3.65 | 2752.0 | 4.95 | 2.3 | 4.87 - 5.03 | 370.3* |
| Incomplete Primary | 2.43 | 1.6 | 2.37 - 2.48 |  | 3.69 | 1.8 | 3.54 - 3.83 |  |
| Complete Primary | 1.96 | 1.4 | 1.92 - 2.01 |  | 3.24 | 1.6 | 3.1 - 3.38 |  |
| Lower Secondary | 1.6 | 1.1 | 1.56 - 1.64 |  | 2.57 | 1.2 | 2.42 - 2.72 |  |
| Upper Secondary | 1.43 | 1 | 1.39 - 1.47 |  | 2.49 | 1.1 | 2.33 - 2.65 |  |
| Post-Secondary | 1.27 | 0.9 | 1.22 - 1.31 |  | 2.08 | 0.9 | 1.95 - 2.2 |  |
| Household Wealth Status |  |  |  |  |  |  |  |  |
| Poorest | 3.4 | 2.4 | 3.34 - 3.46 | 2752.0 | 5.54 | 2.4 | 5.41 - 5.67 | 297.7*** |
| Poorer | 3 | 2.1 | 2.95 - 3.05 |  | 4.82 | 2.3 | 4.7 - 4.95 |  |
| Middle | 2.83 | 2.1 | 2.77 - 2.88 |  | 4.63 | 2.2 | 4.51 - 4.75 |  |
| Richer | 2.6 | 1.9 | 2.55 - 2.65 |  | 4.29 | 2.2 | 4.15 - 4.43 |  |
| Richest | 2.27 | 1.6 | 2.21 - 2.32 |  | 3.42 | 1.9 | 3.29 - 3.54 |  |
| Unmet Need of Family Planning |  |  |  |  |  |  |  |  |
| Unmet Need | 2.76 | 2.2 | 2.69 - 2.83 | 11.3 | 5.37 | 2.7 | 5.19 - 5.55 | 283.0<br>*** |
| Met Need | 2.83 | 2 | 2.79 - 2.86 |  | 4.36 | 2.2 | 4.28 - 4.44 |  |
| Women's Empowerment Index |  |  |  |  |  |  |  |  |
| Not Making Any Decision | 2.89 | 2.2 | 2.85 - 2.93 | 84.5 | 4.88 | 2.4 | 4.78 - 4.97 | 269.2<br>*** |
| Partial Decision Making | 2.65 | 1.8 | 2.61 - 2.7 |  | 3.95 | 2 | 3.86 - 4.04 |  |
| Decision Making Alone | 2.68 | 1.6 | 2.58 - 2.78 |  | 3.52 | 1.9 | 3.28 - 3.76 |  |
| Notes: SD = Standard deviation. Data are from the author's computation from the pooled six-round NDHS dataset (1996 – 2022) |  |  |  |  |  |  |  |  |
| ***p< 0.001; **p < 0.01; *p < 0.05 |  |  |  |  |  |  |  |  |

### Determinants of Fertility Heterogeneity: Poisson Regression Model

The results from the Poisson regression model analysing lifetime fertility are presented in **Error! Reference source not found.** and **Error! Reference source not found.**. The table illustrates the net effect of predictor variables in terms of Incidence Rate Ratios (IRR) and Confidence Intervals (CI) for the expected number of CEB among currently married women, for both lifetime and completed fertility. The main findings are summarised below.

**Table 5.**
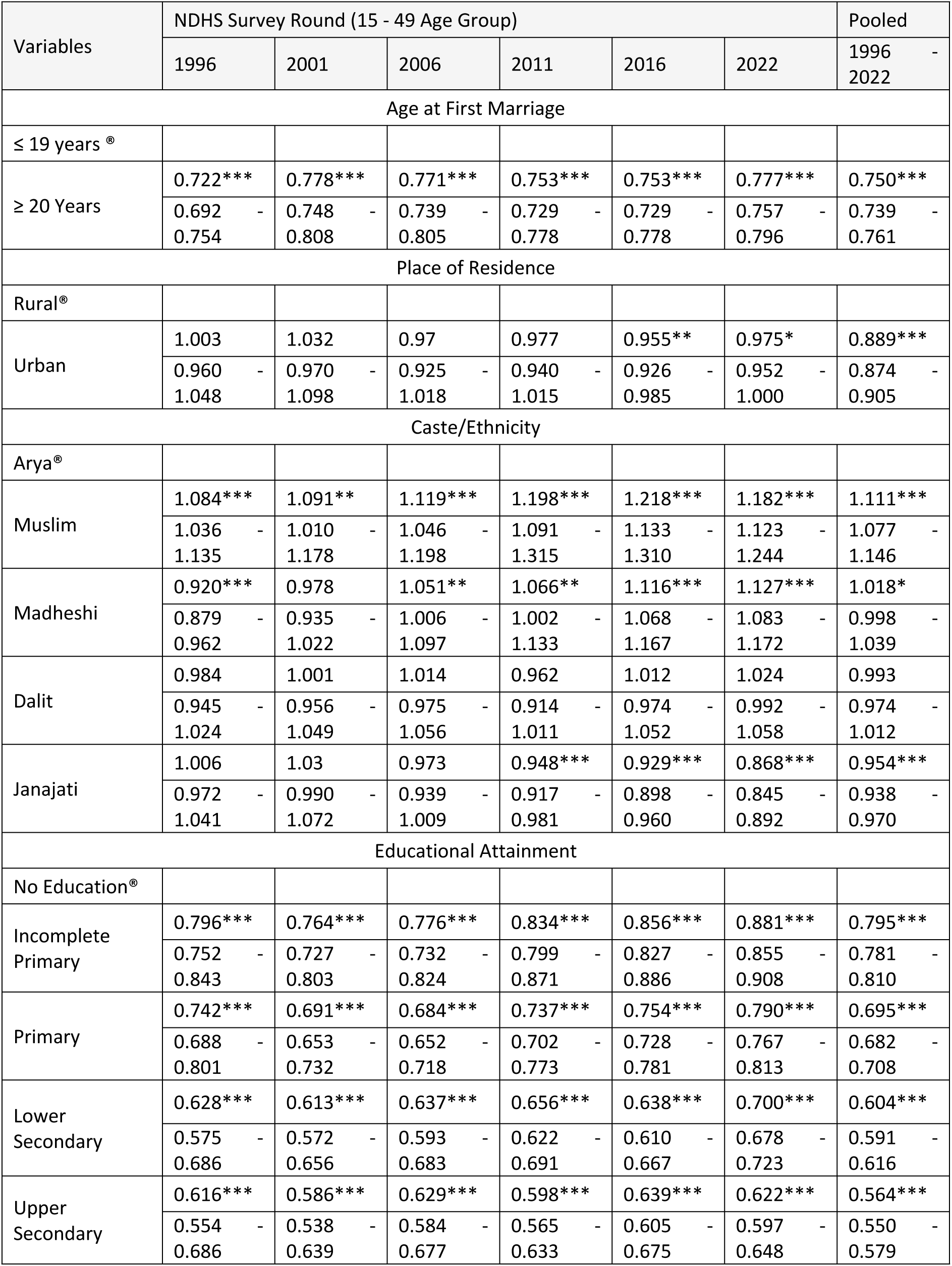
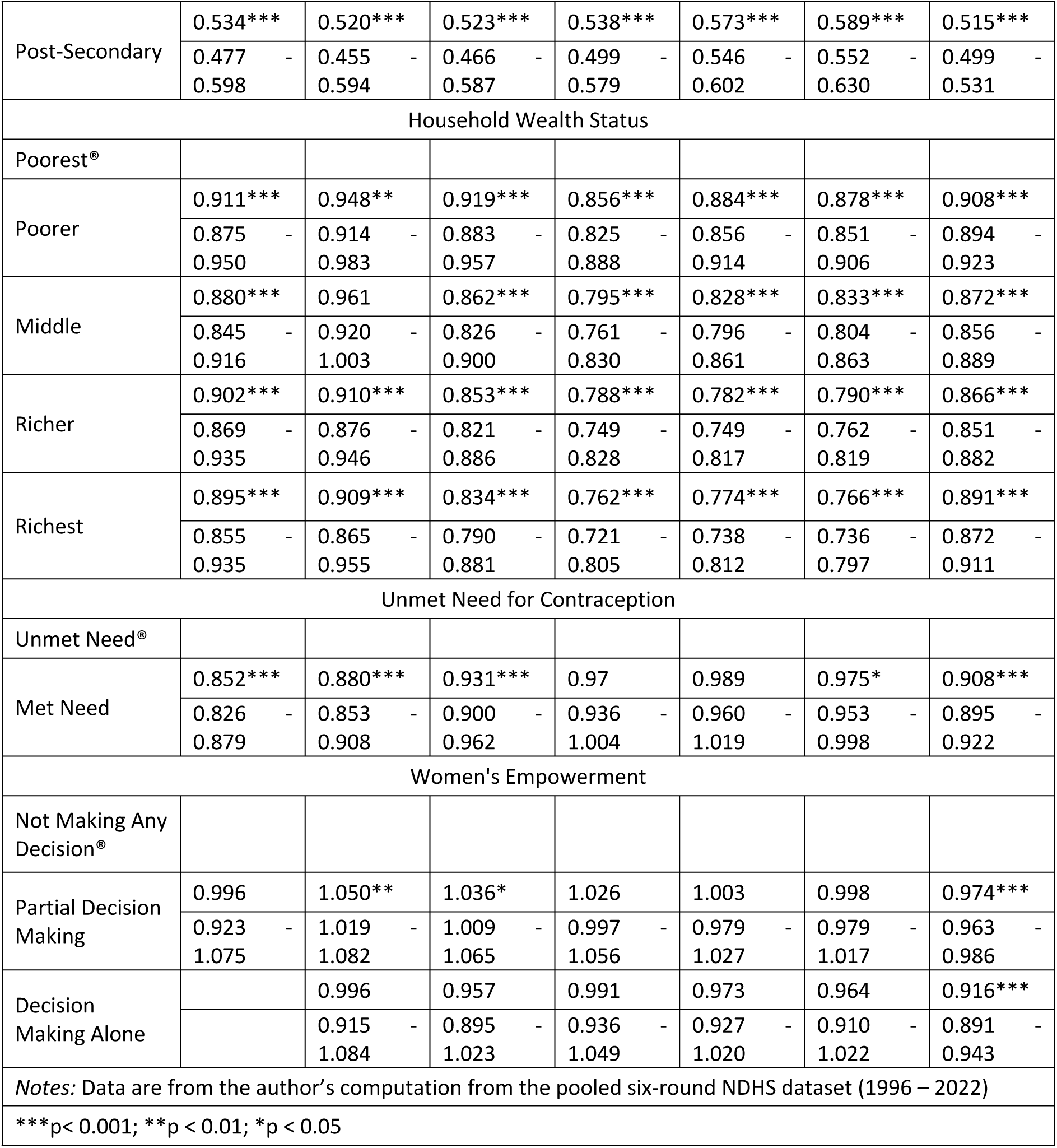
Multivariate Poisson regression model of individual-level factors associated with total number of children ever born with currently married women of reproductive age (15-49 years) in Nepal, using data from 1996-2022 NDHS.

| Variables | NDHS Survey Round (15 - 49 Age Group) |  |  |  |  |  | Pooled |
| --- | --- | --- | --- | --- | --- | --- | --- |
|  | 1996 | 2001 | 2006 | 2011 | 2016 | 2022 | 1996 - 2022 |
| Age at First Marriage |  |  |  |  |  |  |  |
| ≤ 19 years<sup>®</sup> |  |  |  |  |  |  |  |
| ≥ 20 Years | 0.722*** | 0.778*** | 0.771*** | 0.753*** | 0.753*** | 0.777*** | 0.750*** |
|  | 0.692 - | 0.748 - | 0.739 - | 0.729 - | 0.729 - | 0.757 - | 0.739 - |
|  | 0.754 | 0.808 | 0.805 | 0.778 | 0.778 | 0.796 | 0.761 |
| Place of Residence |  |  |  |  |  |  |  |
| Rural<sup>®</sup> |  |  |  |  |  |  |  |
| Urban | 1.003 | 1.032 | 0.97 | 0.977 | 0.955** | 0.975* | 0.889*** |
|  | 0.960 - | 0.970 - | 0.925 - | 0.940 - | 0.926 - | 0.952 - | 0.874 - |
|  | 1.048 | 1.098 | 1.018 | 1.015 | 0.985 | 1.000 | 0.905 |
| Caste/Ethnicity |  |  |  |  |  |  |  |
| Arya<sup>®</sup> |  |  |  |  |  |  |  |
| Muslim | 1.084*** | 1.091** | 1.119*** | 1.198*** | 1.218*** | 1.182*** | 1.111*** |
|  | 1.036 - | 1.010 - | 1.046 - | 1.091 - | 1.133 - | 1.123 - | 1.077 - |
|  | 1.135 | 1.178 | 1.198 | 1.315 | 1.310 | 1.244 | 1.146 |
| Madheshi | 0.920*** | 0.978 | 1.051** | 1.066** | 1.116*** | 1.127*** | 1.018* |
|  | 0.879 - | 0.935 - | 1.006 - | 1.002 - | 1.068 - | 1.083 - | 0.998 - |
|  | 0.962 | 1.022 | 1.097 | 1.133 | 1.167 | 1.172 | 1.039 |
| Dalit | 0.984 | 1.001 | 1.014 | 0.962 | 1.012 | 1.024 | 0.993 |
|  | 0.945 - | 0.956 - | 0.975 - | 0.914 - | 0.974 - | 0.992 - | 0.974 - |
|  | 1.024 | 1.049 | 1.056 | 1.011 | 1.052 | 1.058 | 1.012 |
| Janajati | 1.006 | 1.03 | 0.973 | 0.948*** | 0.929*** | 0.868*** | 0.954*** |
|  | 0.972 - | 0.990 - | 0.939 - | 0.917 - | 0.898 - | 0.845 - | 0.938 - |
|  | 1.041 | 1.072 | 1.009 | 0.981 | 0.960 | 0.892 | 0.970 |
| Educational Attainment |  |  |  |  |  |  |  |
| No Education<sup>®</sup> |  |  |  |  |  |  |  |
| Incomplete Primary | 0.796*** | 0.764*** | 0.776*** | 0.834*** | 0.856*** | 0.881*** | 0.795*** |
|  | 0.752 - | 0.727 - | 0.732 - | 0.799 - | 0.827 - | 0.855 - | 0.781 - |
|  | 0.843 | 0.803 | 0.824 | 0.871 | 0.886 | 0.908 | 0.810 |
| Primary | 0.742*** | 0.691*** | 0.684*** | 0.737*** | 0.754*** | 0.790*** | 0.695*** |
|  | 0.688 - | 0.653 - | 0.652 - | 0.702 - | 0.728 - | 0.767 - | 0.682 - |
|  | 0.801 | 0.732 | 0.718 | 0.773 | 0.781 | 0.813 | 0.708 |
| Lower Secondary | 0.628*** | 0.613*** | 0.637*** | 0.656*** | 0.638*** | 0.700*** | 0.604*** |
|  | 0.575 - | 0.572 - | 0.593 - | 0.622 - | 0.610 - | 0.678 - | 0.591 - |
|  | 0.686 | 0.656 | 0.683 | 0.691 | 0.667 | 0.723 | 0.616 |
| Upper Secondary | 0.616*** | 0.586*** | 0.629*** | 0.598*** | 0.639*** | 0.622*** | 0.564*** |
|  | 0.554 - | 0.538 - | 0.584 - | 0.565 - | 0.605 - | 0.597 - | 0.550 - |
|  | 0.686 | 0.639 | 0.677 | 0.633 | 0.675 | 0.648 | 0.579 |
| Post-Secondary | 0.534*** | 0.520*** | 0.523*** | 0.538*** | 0.573*** | 0.589*** | 0.515*** |
|  | 0.477 - | 0.455 - | 0.466 - | 0.499 - | 0.546 - | 0.552 - | 0.499 - |
|  | 0.598 | 0.594 | 0.587 | 0.579 | 0.602 | 0.630 | 0.531 |
| Household Wealth Status |  |  |  |  |  |  |  |
| Poorest® |  |  |  |  |  |  |  |
| Poorer | 0.911*** | 0.948** | 0.919*** | 0.856*** | 0.884*** | 0.878*** | 0.908*** |
|  | 0.875 - | 0.914 - | 0.883 - | 0.825 - | 0.856 - | 0.851 - | 0.894 - |
|  | 0.950 | 0.983 | 0.957 | 0.888 | 0.914 | 0.906 | 0.923 |
| Middle | 0.880*** | 0.961 | 0.862*** | 0.795*** | 0.828*** | 0.833*** | 0.872*** |
|  | 0.845 - | 0.920 - | 0.826 - | 0.761 - | 0.796 - | 0.804 - | 0.856 - |
|  | 0.916 | 1.003 | 0.900 | 0.830 | 0.861 | 0.863 | 0.889 |
| Richer | 0.902*** | 0.910*** | 0.853*** | 0.788*** | 0.782*** | 0.790*** | 0.866*** |
|  | 0.869 - | 0.876 - | 0.821 - | 0.749 - | 0.749 - | 0.762 - | 0.851 - |
|  | 0.935 | 0.946 | 0.886 | 0.828 | 0.817 | 0.819 | 0.882 |
| Richest | 0.895*** | 0.909*** | 0.834*** | 0.762*** | 0.774*** | 0.766*** | 0.891*** |
|  | 0.855 - | 0.865 - | 0.790 - | 0.721 - | 0.738 - | 0.736 - | 0.872 - |
|  | 0.935 | 0.955 | 0.881 | 0.805 | 0.812 | 0.797 | 0.911 |
| Unmet Need for Contraception |  |  |  |  |  |  |  |
| Unmet Need® |  |  |  |  |  |  |  |
| Met Need | 0.852*** | 0.880*** | 0.931*** | 0.97 | 0.989 | 0.975* | 0.908*** |
|  | 0.826 - | 0.853 - | 0.900 - | 0.936 - | 0.960 - | 0.953 - | 0.895 - |
|  | 0.879 | 0.908 | 0.962 | 1.004 | 1.019 | 0.998 | 0.922 |
| Women's Empowerment |  |  |  |  |  |  |  |
| Not Making Any Decision® |  |  |  |  |  |  |  |
| Partial Decision Making | 0.996 | 1.050** | 1.036* | 1.026 | 1.003 | 0.998 | 0.974*** |
|  | 0.923 - | 1.019 - | 1.009 - | 0.997 - | 0.979 - | 0.979 - | 0.963 - |
|  | 1.075 | 1.082 | 1.065 | 1.056 | 1.027 | 1.017 | 0.986 |
| Decision Making Alone |  | 0.996 | 0.957 | 0.991 | 0.973 | 0.964 | 0.916*** |
|  |  | 0.915 - | 0.895 - | 0.936 - | 0.927 - | 0.910 - | 0.891 - |
|  |  | 1.084 | 1.023 | 1.049 | 1.020 | 1.022 | 0.943 |
| Notes: Data are from the author's computation from the pooled six-round NDHS dataset (1996 – 2022) |  |  |  |  |  |  |  |
| ***p< 0.001; **p < 0.01; *p < 0.05 |  |  |  |  |  |  |  |

**Table 6.**
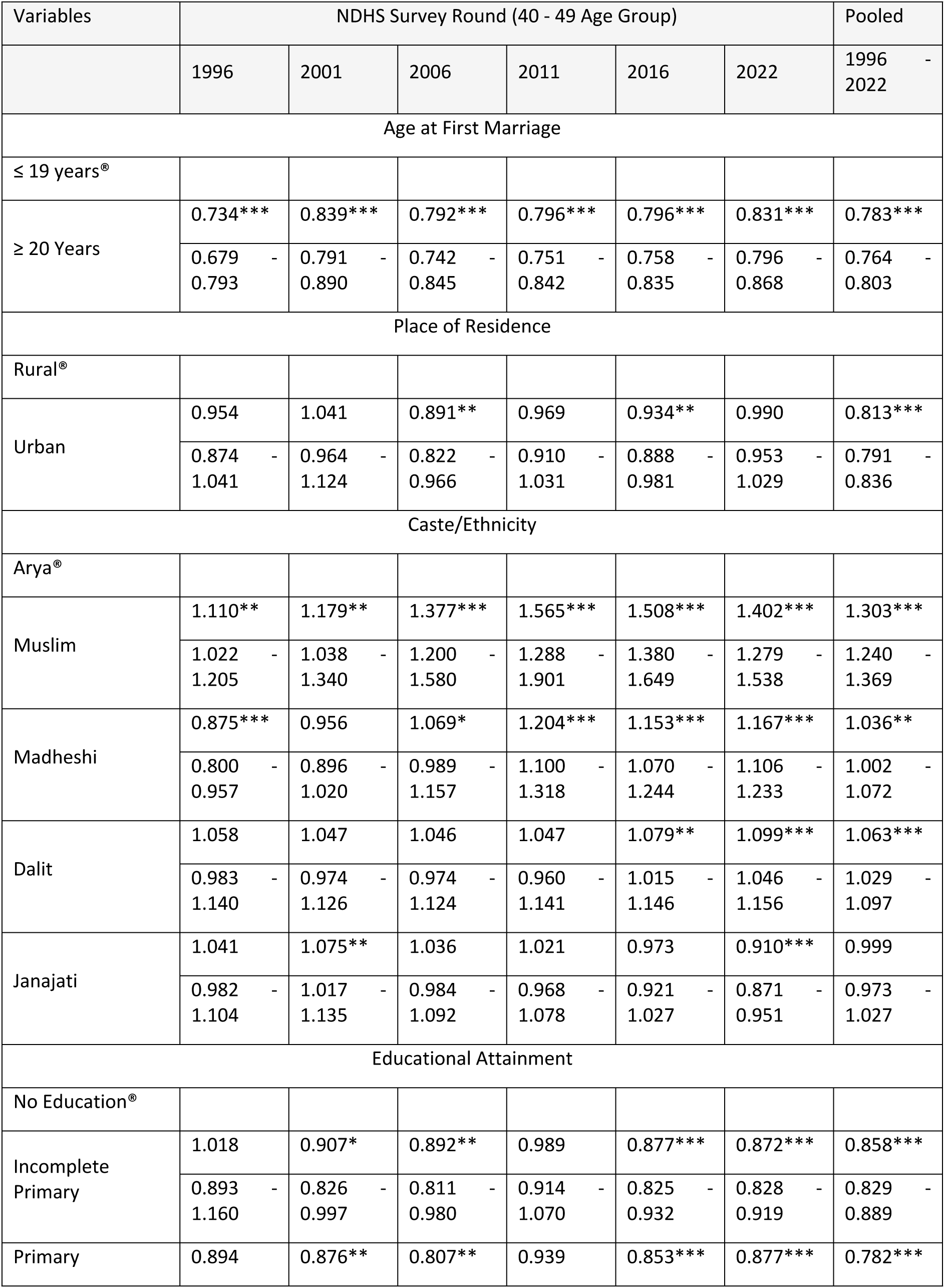
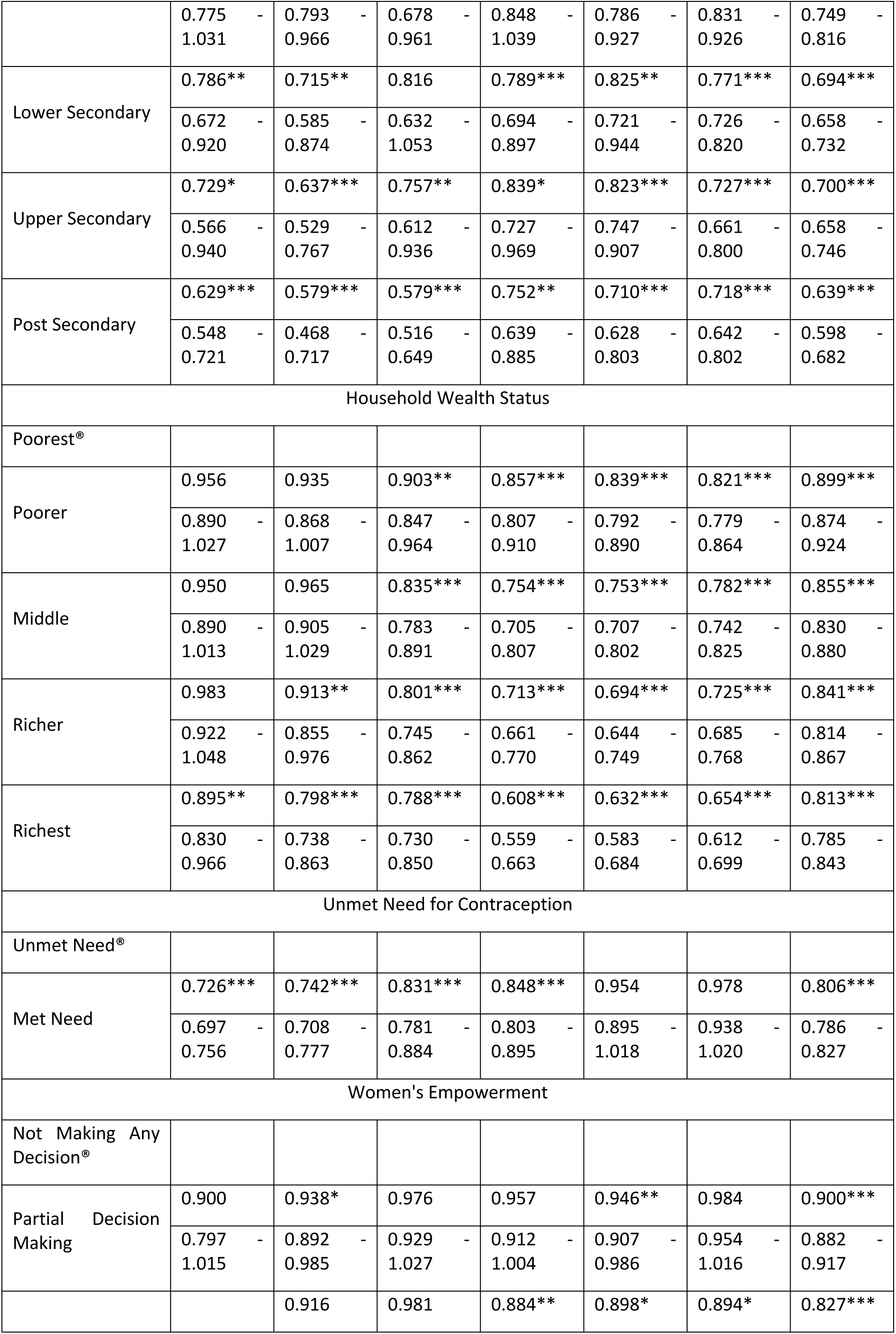
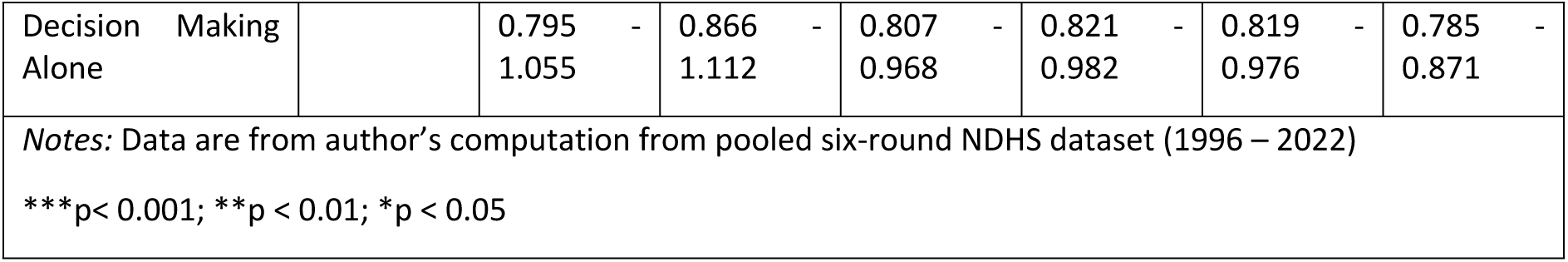
Multivariate Poisson regression model of individual-level factors associated with total number of children ever born with currently married women of reproductive age (15-49 years) in Nepal, using data from 1996-2022 NDHS.

### Age at first marriage

The variable is significantly associated with the number of CEB across all NDHS rounds and the pooled dataset (*p<0.001*) for both lifetime and completed fertility model. For the pooled data period:

Lifetime fertility: Woman who married at age 20 or older had 25% fewer expected children (IRR=0.75; 95% CI 0.74-0.76) compared to those who married at age 19 or younger, holding other predictors constant, and

Completed fertility: Woman who married at age 20 or older had 21.7% fewer expected children (IRR=0.78; 95% CI 0.76-0.80) compared to those who married at age 19 or younger, holding other predictors constant.

Individual NDHS round analysis: There was a consistent pattern of lower fertility with late age at first marriage was observed across the NDHS round for both lifetime and completed fertility as follows.

Lifetime fertility: Woman who married at age 20 or older had 27.8% fewer expected children in NDHS 1996 (IRR = 0.72, 95% CI 0.69-0.75), 22.2% fewer in NDHS 2001 (IRR = 0.78, 95% CI 0.75-0.81), 22.9% fewer in 2006 (IRR = 0.77, 95% CI 0.74-0.80), 24.7% fewer in both NDHS 2011 and 2016 (IRR = 0.75, 95% CI 0.73-0.78), and 22.3% fewer in 2022 NDHS (IRR = 0.78, 95% CI 0.76-0.80) compared to those who married at 19 or younger, holding other predictors constant.

Completed fertility: Women who married at age 20 or older had fewer expected children ranging from 26.6% (IRR = 0.73, 95% CI 0.68-0.79) in 1996 NDHS to 16.9% (IRR=0.83, 95% CI 0.80-0.87) in 2022 NDHS compared to the reference category of ≤ 19 years of age at first marriage, holding other predictors constant.

The contracting range of expected CEB over the six rounds of NDHS suggests a significant role in changing social norms. This phenomenon contributes to a continued reduction in fertility, ever women married before 20 years of age, in an environment of higher prevalence of women with late marriage.

### Place of residence

The place of residence was significantly associated with the number of CEB for the pooled data period in both the lifetime and completed fertility models as follows:

Woman living in urban areas had 11.1% fewer expected children (IRR = 0.89, 95% CI 0.87-0.90) compared to those living in rural areas (*p<0.001*), holding other predictors constant.

Women living in urban areas had 18.7% fewer expected children (IRR = 0.81, 95% CI 0.79-0.84) compared to those living in rural areas (p < *0.001*), holding other predictors constant.

Individual NDHS round analysis: A micro-level examination of NDHS rounds reveals that there was an inconsistent pattern of lower fertility with place of residence and fertility for both lifetime and completed fertility as follows:

Lifetime fertility: The first four NDHS rounds (1996, 2001, 2006, and 2011) were not statistically significant. However, the 2016 (p < 0.01) and 2022 (p < *0.05*) NDHS rounds showed significant associations with place of residence. Women living in urban areas had 4.5% fewer expected number of CEB in NDHS 2016 (IRR = 0.96, 95% CI 0.93-0.99) and 2.5% fewer in NDHS 2022 (IRR = 0.98, 95% CI 0.95-1.00) compared to those living in rural areas, holding other predictors constant.

Completed fertility: Only two rounds of NDHS (2006 and 2016) results were statistically significant (*p<0.05*). Women living in urban areas had 10.9% fewer expected number of CEB in 2006 (IRR=0.89, 95% CI 0.82-0.97) and 6.6% fewer in 2016 (IRR=0.93, 95% CI 0.89-0.98) compared to those living in rural area, holding other predictors constant.

### Caste/ethnicity and religion

The pooled and nearly all NDHS rounds highlighted that not all caste/ethnicities were statistically significant in both lifetime and completed fertility models, reinforcing the complexity of the relationship between caste/ethnicity and CEB, which may vary across time and space. The pooled data analysis is presented below:

Lifetime fertility: Muslim women had 11.1% higher expected children (IRR=1.11; 95% CI 1.08-1.15), and Madheshi women had 1.8% higher expected children (IRR=1.02; 95% CI 1.0-1.04), while Dalit had 4.6% fewer expected children (IRR=1.06; 95% CI 1.03-1.1) compared to the Arya women. In contrast, Dalit women showed no significant difference compared to Arya women. Janajati women had 5% fewer expected children (IRR = 0.94; 95% CI 0.94 - 0.97).

Completed Fertility: The Muslim, Dalit, and Madheshi women had 30%, 6%, and 4% higher expected children as compared to Arya women. In contrast, Janajati women showed no significant difference (IRR=0.99; 95% CI 0.97-1.03) compared to Arya women.

Individual NDHS round analysis: This analysis indicated shifting trends in the CEB among the studied caste/ethnicities compared to Arya women in both lifetime and completed fertility as follows:

Lifetime fertility: Muslim women had 8.4%, 9.1%, 11.9%, 19.8%, 21.8%, and 18.2% higher CEB than Arya women in the 1996, 2001, 2006, 2011, 2016, and 2022 NDHS rounds, respectively. Janajati women had 0.6% and 3.0% higher CEB in the 1996 and 2001 NDHS rounds, respectively, but lower CEB by 2.7%, 5.2%, 7.1% and 13.2% in the 2006, 2011, 2016 and 2022 NDHS rounds, respectively, compared to Arya women.

Interestingly, Madheshi women initially had lower CEB in the 1996 and 2001 NDHS rounds, with 8.0% and 2.2% fewer CEB, respectively, compared to Arya women. However, in the subsequent 2006, 2011, 2016, and 2022 NDHS rounds, Madheshi women had higher CEB by 5.1%, 6.6%, 11.6%, and 12.7%, respectively, compared to Arya women. In the case of Dalit women, there was no significant association compared to Arya women.

Completed fertility: It shows a similar pattern of results to the lifetime fertility model. However, nuanced and shifting trends emerged across the six NDHS rounds. This may be the impact of changes in social norms, an increase in educational attainment, the use of contraceptives, male labour migration, and increased use of information technology.

### Educational attainment

Across all NDHS rounds and the pooled data period, women with different educational levels had significantly lower CEB compared to those with no education (p < *0.001*). The reductions in CEB for the pooled data period in both lifetime and completed fertility models are presented below:

Lifetime fertility: The reductions in CEB were: incomplete primary (IRR = 0.80, 20.5% reduction, 95% CI 0.78–0.81), complete primary (IRR = 0.70, 30.5% reduction, 95% CI 0.68-0.71), lower secondary (IRR = 0.60, 39.6% reduction, 95% CI 0.59-0.62), upper secondary (IRR = 0.56, 43.6% reduction, 95% CI 0.55-0.58), and post-secondary (IRR = 0.51, 48.5% reduction, 95% CI 0.50-0.53) compared to women with no education.

Completed fertility: The reductions in CEB with different education levels were: incomplete primary (IRR = 0.86, 14.2% reduction, 95% CI 0.83–0.89), complete primary (IRR = 0.78, 21.8% reduction, 95% CI 0.75-0.82), lower secondary (IRR = 0.69, 30.6% reduction, 95% CI 0.66-0.73), upper secondary (IRR = 0.70, 30.0% reduction, 95% CI 0.66-0.75), and post-secondary (IRR = 0.64, 36.1% reduction, 95% CI 0.60-0.68) compared to the women with no education.

#### Individual NDHS round analysis

Lifetime fertility: Consistent lower fertility was observed across educational attainment levels. The range of expected CEB reduction for educated women (incomplete primary to upper secondary) was NDHS 1996 (20.4% - 46.6%), NDHS 2001 (23.6%-48%), NDHS 2006 (22.4%-47.7%), NDHS 2011 (16.6%-46.2%), NDHS 2016 (14.4-42.7%, and NDHS 2022 (11.9%-41.1%) compared to women with no education. The expanding range of expected CEB over the six rounds of NDHS (1996–2022) indicates significant educational diffusion effects, contributing to continued fertility reduction, even for women with lower education levels.

Completed fertility: In the first four NDHS rounds, the significance of incomplete and complete primary education was not observed due to the presumably low educational attainment. However, secondary and higher education had a strong adverse effect on fertility. In the last two NDHS rounds, the expected CEB reduction for educated women (incomplete primary to upper secondary) was 12.3% to 29.0% in NDHS 2016 and 12.8% to 28.2% in NDHS 2022, compared to women with no education, holding other predictors constant. Education remains highly significant and consistent at the individual level, with more substantial negative effects on fertility for secondary and higher education.

### Household socioeconomic wealth status

Across all individual NDHS rounds and the pooled data period, women from wealthier households had significantly lower CEB compared to those from poorer households (*p < 0.001*). For the pooled data period:

Lifetime fertility: The reductions in CEB were: poorer households (IRR = 0.91, 9.2% reduction, 95% CI 0.89–0.92), middle (IRR = 0.87,12.8% reduction, CI 0.86-0.89), richer (IRR = 0.87, 13.4% reduction, CI 0.85-0.88), and richest (IRR = 0.89, 10.9% reduction, 95% CI 0.87-0.91) compared to the poor households, holding other predictor constant.

Completed fertility: Households with different wealth status exhibited varying reduction rates: poorer households (IRR = 0.90, 10.1% reduction, CI 0.87–0.92), middle (IRR = 0.85, 14.5% reduction, CI 0.83-0.88), richer (IRR = 0.84, 15.9% reduction, CI 0.81-0.87), and richest (IRR = 0.81, 18.7% reduction, 95% CI 0.78-0.84) compared to the poor households, holding other predictor constant.

Individual NDHS round analysis: Inconsistent and varying results were observed across the wealth quintiles, indicating a reduction in fertility.

Lifetime fertility: The range of expected CEB reduction for poorer to rich households was: NDHS 1996 (8.9% - 12.0%), followed by NDHS 2001 (3.9%-9.1%), NDHS 2006 (8.1%-16.6%), NDHS 2011 (14.4%-23.8%), NDHS 2016 (11.6%-22.8%), and NDHS 2022 (12.2%-23.4%) compared to women from poor households, holding other predictor variables constant.

Completed fertility: In the first two NDHS rounds, inconsistent and varying results were observed. In the 1996 NDHS, only the richest households were found to be statistically significant (p < 0.01), with women living in the richest households having 10.5% fewer expected CEBs (IRR = 0.90, 95% CI 0.83-0.97) compared to women from poor households, holding other predictors constant. In the 2001 NDHS, only richer and richest households were statistically significant, with women in these households having 8.7% (IRR=0.91, 95% CI 0.85-0.98) and 20.2% (IRR=0.80, 95% CI 0.74-0.86) fewer expected CEB, respectively compared to the women from poor households, holding other predictors constant. The remaining three NDHS rounds showed a consistent pattern of lower CEB among households from poorer to richest compared to the poorest households (p < *0.001)*.

### Unmet need for contraception

In the pooled data analysis period, women with met contraception needs had significantly fewer CEB compared to those with unmet needs (*p<0.001*).

Lifetime fertility: Women with a met need for contraception had 9.2% fewer expected children (IRR = 0.91, 95% CI 0.90-0.92) compared to those with unmet needs for contraception, holding other variables constant.

Completed fertility: Women with met need of contraception had 19.4% fewer expected children (IRR = 0.81, 95% CI 0.79-0.83) compared to those with unmet need for contraception, holding other variables constant.

Individual NDHS round analysis: There was no consistent pattern of effect on unmet need for contraception at individual NDHS rounds for both lifetime and completed fertility models.

Lifetime fertility: Significant reductions in CEB were observed in the NDHS 1996, 2001, and 2006 (p < 0.001), and in the NDHS 2022 (p < *0.05*). No significant effect was observed in the NDHS surveys of 2011 and 2016. The analysis revealed a decreasing trend in the effect of unmet need for contraception, ranging from NDHS 1996 (IRR = 0.85, 14.8% reduction, 95% CI 0.83-0.88) to NDHS 2022 (IRR = 0.97, 2.5% reduction, 95% CI 0.95-0.99).

Completed fertility: There was no consistent pattern of effect on unmet need for contraception. In the first four NDHS rounds (1996, 2001, 2006, and 2011), a significantly lower number of CEB was observed (p<0.001). The analysis revealed a decreasing trend in the effect of unmet need for contraception, ranging from NDHS 1996 (IRR = 0.73, 27.4% reduction, 95% CI 0.70-0.76) to NDHS 2011 (IRR = 0.85, 15.2% reduction, 95% CI 0.80-0.89).

In contrast, no statistically significant results were observed for unmet need for contraception in the 2016 and 2022 NDHS rounds.

#### Women’s empowerment

Women with decision-making capacity were significantly associated with the number of CEB across all NDHS rounds and the pooled dataset (p < 0.001) for both lifetime and completed fertility models. The results from the pooled data period have been presented as follows:

Lifetime fertility: Women with partial decision-making had 2.6% fewer expected children (IRR = 0.97, 95% CI 0.96-0.99), and those with full decision-making had 8.4% fewer expected children (IRR = 0.92, 95% CI 0.89-0.94) compared to those without decision-making capacity, holding other predictors constant.

Completed fertility: Women with partial decision-making had 10.0% fewer expected children (IRR = 0.90, 95% CI 0.88-0.92), and those with full decision-making had 17.3% fewer expected children (IRR = 0.83, 95% CI 0.76-0.87) compared to those without decision-making capacity, holding other predictors constant.

Individual NDHS round analysis: Women with varying levels of empowerment exhibited different fertility rates in both lifetime and completed fertility models.

Lifetime fertility: Out of six NDHS rounds, only NDHS 2001 (IRR = 1.05; 5% increase, 95% CI 1.05-1.08) and NDHS 2006 (IRR = 1.04; 3.6% increment, 95% CI 1.01-1.07) were statistically significant at p<0.001 and p<0.05, respectively, for women with partial decision making compared to the women with no decision-making capacity. These findings suggest that more empowered women having lower fertility compared to less empowered women.

Completed fertility: Out of six NDHS rounds, only NDHS 2001 (p<0.05) and 2016 (p<0.01) showed statistically significant results for women with partial decision-making. In these rounds, women with partial decision-making had 6.2% and 5.4% fewer expected children, respectively, compared to women with no decision-making capacity. Additionally, the last three NDHS rounds showed statistically significant results, with women in the decision-making alone category approximately 10.8% fewer expected CEB compared to women with no decision-making, holding other predictors constant. These robust findings suggest that the relationship between women’s empowerment and CEB is complex and may vary across time and space.

## Discussions

The primary objective of this study was to investigate the heterogeneity of fertility in Nepal. Various studies debating the relative roles of urbanisation, education, and family planning programmes in declining fertility in Nepal are inconclusive. This paper examined whether caste/ethnicity and educational attainment, along with associated covariates of the women, were the primary sources of heterogeneous fertility. The outcome variable was the total number of CEB. Data on currently married women of reproductive age (15-49 years) at the time of the survey were analysed using Poisson regression models.

### Age at first marriage

was found to be a significant determinant of the number of CEB (96). Early marriage leads to a longer exposure period and higher fertility, while late marriage has a fertility-reducing effect. Child marriage, defined as marriage before the age of 18, has been illegal in Nepal since 1963. The current law sets the minimum age for marriage at 20 for both men and women (97). Despite this, Nepal faces significant challenges with child marriage(98). Multivariate analysis of individual NDHS rounds and pooled data (1996-2022) showed that age at marriage is statistically significant for both lifetime and completed fertility. This result is consistent with studies in developing countries (99), Asian countries (100,101), Sub-Saharan African countries (102,103), and Latin American countries (104).

### Place of residence

Over the long term, place of residence has been significantly associated with fertility patterns for both lifetime and completed in Nepal, as evidenced by the pooled data from 1996-2022 (*p<0.01*). Urban areas consistently exhibited lower fertility rates compared to rural regions, attributed to factors such as population concentration, individualisation, anonymisation, cultural heterogeneity and regimentation (105) This finding is consistent with studies in the Global South (106), India (107), Bangladesh (25), Myanmar (108), and Thailand (109). However, the effects of urbanisation on fertility were not always statistically significant or negligible.

The inconsistency can partly be explained by the evolving nature of urbanisation in Nepal. Urban and peri-urban areas are combined into a single category in the NDHS classification, which obscures the distinctive characteristics of fully urbanised settings. According to the National Statistics Office (110), Nepal’s population is currently distributed as 27.1% urban, 39.7% peri-urban, and 33.2% rural. Nepal comprises 293 municipalities and 460 rural municipalities across its seven provinces. Rapid urban expansion beyond official boundaries and changes in administrative definitions make it challenging to measure urbanisation levels and their impact on fertility accurately. Understanding the diffusion of fertility decline from urban to rural areas remains critical to comprehending population changes in Nepal’s diverse geographic and sociocultural landscapes.

### Caste/Ethnicity and Religion

Over the long term, caste/ethnicity was significantly associated with the number of children ever born (CEB) in both lifetime and completed fertility. However, there was no consistent pattern of statistical significance across the six rounds of the NDHS for different castes and ethnicities. For example, Dalit women showed a fluctuating trend of decreasing expected CEB by 9.3% in 1996, followed by 8.2% in 2001, 9.4% in 2006, 19.7% in 2011, 16.9% in 2016, and 13.3% in 2022 compared to Muslim women, holding other predictors constant. Not all caste or ethnicities were statistically significant, reinforcing the presumption that caste or ethnicity has varying levels of impact.

### Women’s educational attainment

Women’s education level was significantly associated with the number of CEB. This finding is consistent with studies in developing countries (111), Sub-Saharan Africa (110), and other South Asian countries, which have found that secondary or higher education levels are inversely and significantly associated with the number of CEB. This phenomenon has been observed in India (107,112), Bangladesh (25), and Pakistan (113).

Educated women are more likely to be informed about reproductive health issues, resulting in fewer number of CEB (32,114–117) . Women’s empowerment in the household could moderate the nexus between low education levels and fertility. Secondary or higher education enhances cognitive abilities, enabling women to participate, reflect, and act on their life conditions, access knowledge, and make informed decisions, thereby claiming their empowerment (118,119). Studies in Nepal (53,55), Timor-Leste (64), Gambia (62), Nigeria (120), and Burkina Faso (121) found a decrease in CEB among women with secondary or higher education. From the analysis, fertility patterns mostly differ by educational attainment rather than other factors such as place of residence, caste/ethnicity, and unmet need for contraception.

### Household economic status

Household economic status was found to be a determinant of the number of CEB. Women from rich and middle-income households were inversely and significantly associated with the number of CEB. This finding is consistent with studies in other South Asian countries, such as Bangladesh (59, 95), Kenya (122) and Ghana and Nigeria (123). It also aligns with finding in Nepal (124,125). Women’s empowerment in the household can moderate the association between household economic status and fertility.

Increased economic status is highly correlated with exposure to mass media, as wealthier households can afford radio and television sets (126). This exposure helps women to access information on reproductive health and human rights, freeing them from traditional norms and empowering them within the household compared to women from poor households.

### Unmet Need for Contraception

Over the long term, unmet need for contraception was found to be a determinant of the number of CEB for both lifetime and completed fertility. However, not all individual NDHS rounds showed significant results, reinforcing the presumption that the unmet need for contraception has varying levels of impact. There was no consistent pattern of effect across individual NDHS rounds.

Similar studies have highlighted the complex relationship between the unmet need for contraception and fertility. For instance, Machiyama et al. (2017) emphasised the gap between women’s reproductive desires and contraceptive behaviour (127). Anik et al. (2022) examined the socioeconomic factors influencing the unmet need for modern contraception in low- and lower-middle-income countries (128). A scoping review by Sedgh et al. (2016) discussed the determinants of unmet need for family planning (129). Bradley et al. (2012) explored the impact of unmet need for contraception on fertility preferences and outcomes (130). Casterline and Sinding (2000) provided insights into the reasons behind the unmet need for family planning and its implications for fertility (131).

### Women’s empowerment

Over the long term, women’s empowerment was significantly associated with the number of CEB. Empowered women tend to make rational decisions, including fertility decisions, and are likely to opt for fewer children. It can be deduced that living in urban areas, having at least secondary education and being from middle or rich households moderate women’s empowerment in the household, which in turn affects fertility.

In individual NDHS rounds, not all levels of women’s empowerment were significant, reinforcing the presumption that empowerment has varying levels of impact. The results indicate that empowerment may reduce fertility levels, particularly if it affects not only fertility decisions but also preferences for boys over girls. Similar results were found in (57).

## Conclusions and Recommendations

The results of the Poisson regression model provide robust confirmation of fertility heterogeneity. The study concludes that several factors affect the heterogeneity of fertility. Age at first marriage is more than a personal milestone; it’s a biological variable with a decisive impact on the expected number of children. The results provide statistical clarity on the relationship between age at first marriage and both lifetime and completed fertility. Regarding place of residence, the findings highlight the dynamic nature of the relationship between residence and the number of CEB, emphasising the need for further research on male labour migration abroad, the practice of induced abortion in rural areas, peri-urban fertility behaviours, etc.

Caste/ethnicity and religion also significantly affect the number of CEB in both lifetime and completed fertility. However, there was an inconsistent pattern of statistically significant results, with a fluctuating trend in the expected number of CEB compared to Arya women. These shifting trends across six NDHS rounds suggest that the relationship between caste and ethnicity may vary over time and space, influenced by changes in social norms, the diffusion of modern contraception, male labour migration, and educational diffusion effects. Similarly, although statistically significant results were obtained for the unmet need for contraception and women’s empowerment over the long term, there was an inconsistent pattern in individual NDHS rounds, with varying effects on the number of CEB. For example, the impact of both variables was negligible in the last NDHS rounds.

Women’s education has had the most significant impact on the decline in fertility desires compared to other predictor variables. The proportion of educated women in the community influences the decline of fertility among other education groups, implying a diffusion effect where women with lower education learn from more educated women, influenced by the local environment and cultural atmosphere. Over time, this diffusion effect can also contribute to the declining fertility trend among less educated women. Moreover, the multiplier effect of women’s education is linked to informed family planning choices, health care utilisation, and women’s empowerment, resulting in further fertility decline. In conclusion, the statistical analyses indicate that women’s education is the key factor of fertility heterogeneity.

The study recommends that the Government of Nepal invest in women’s education, such as compulsory completion of secondary education. Additionally, the government should emphasise the implementation, monitoring, and evaluation of gender policies at all levels (national and subnational) to ensure women’s empowerment, positively affecting community, institutional, and household levels by reducing women-burdening cultural values and increasing labour force participation rates. Furthermore, the government should strengthen public healthcare programs, including family planning initiatives.

### Strengths and limitations of the study

The strength of this study lies in the six rounds of NDHS datasets, which collect nationally representative household survey data, making the generalisation of the findings more reliable. Additionally, the use of the Poisson regression model is well-suited for outcome variables with count data, representing social phenomena that can be counted. It provides reliable results concerning the predictor variables and the number of CEB. However, there are some limitations associated with the study:

*Self-reported data:* The self-reported nature of the data may influence the results due to the possibility of social desirability bias inherent in respondents’ self-reporting. However, the NDHS has taken measures to collect accurate information and evaluate the quality of the reported data.

*Place of residence*: The delineation of urban and rural areas often has ambiguous boundaries, resulting in regions with mixed characteristics of both urban and rural environments. Additionally, the newly delineated urban areas in Nepal lack adequate and standardised criteria to define urban areas and do not meet generally accepted criteria for urban settlements, including transportation, parks and recreation, services, economic basis, water, and wastewater infrastructure. Furthermore, population size is a key indicator for declaring an area as urban in Nepal, but this criterion alone may not capture all aspects of urbanisation.

*Women’s empowerment:* Another limitation is that women’s empowerment is a complex phenomenon, even within a household context. There is no consensus on a definition or the most important dimensions of women’s empowerment. Thus, women’s empowerment may not have an independent effect on CEB as it may interact with prevailing sociocultural norms. Since we do not have a robust, measurable indicator for reflecting sociocultural norms, we have not tested for the interaction effect.

## Data Availability

Data will be available upon the request from the corresponding Author

https://dhsprogram.com/Countries/Country-Main.cfm?ctry_id=13&c=Nepal&Country=Nepal&cn=&r=4

